# Global Burden and Trends of Childhood and Adolescent Asthma from 1990 to 2023: A Comprehensive Analysis

**DOI:** 10.64898/2026.03.28.26349599

**Authors:** Erhang Yin, Yang Yang, Shirong He, Zexiang Wu, Wei Tan, Fang Du, Chenxi Yang, Haiyang Yu

## Abstract

**Objective:** To comprehensively evaluate the evolution of global childhood and adolescent asthma (ages 0-19) disease burden from 1990-2023, explore spatiotemporal patterns, influencing factors, health equity, and predict future trends.

**Methods:** Using Global Burden of Disease (GBD) data, we analyzed prevalence, incidence, mortality, and disability-adjusted life years (DALYs) rates across global and 21 GBD regions from 1990-2023. Joinpoint regression identified temporal trends, age-period-cohort models analyzed effect contributions, Das Gupta decomposition quantified demographic and epidemiological impacts, inequality indices assessed health equity, and Bayesian models projected 2024-2038 trends.

**Results:** In 2023, the global number of children and adolescents with asthma reached 131 million, with an age-standardized prevalence rate (ASPR) of 1,789.9 per 100,000. From 1990 to 2023, the global ASPR and age-standardized incidence rate (ASIR) of asthma in children and adolescents showed an upward trend, while the age-standardized mortality rate (ASMR) and age-standardized disability-adjusted life years (DALYs) rate (ASDR) exhibited a downward trend. Among the 0-14 age group, the disease burden was greater in males than in females, whereas in the 15-19 age group, males had a lower disease burden than females. Projections indicate that over the next 15 years, the overall disease burden will continue to decline; however, female mortality rates and DALYs rates are projected to show an upward trend.

**Conclusions:** The increasing prevalence and incidence rates, coupled with declining mortality and DALYs rates of asthma among children and adolescents globally, underscore the necessity for targeted public health interventions. These findings provide crucial insights for early diagnosis, treatment optimization, and global health policy formulation.

## 1 Introduction

Asthma is a heterogeneous disease characterized by chronic airway inflammation, reversible airflow limitation, and airway hyperresponsiveness, and has become one of the most common chronic respiratory diseases among children worldwide[1]. According to the Global Burden of Disease Study, approximately 334 million people currently suffer from asthma globally, with children representing the most affected population[2]. Among the 5-14 year age group, asthma ranks among the top 10 causes of disability-adjusted life years globally, not only severely impacting children’s quality of life and physical and mental development, but also imposing a substantial burden on families and healthcare systems[3–5].

In recent years, the Global Burden of Disease Study has provided an important data platform for understanding the epidemiological characteristics of childhood asthma. The Global Asthma Network (GAN) Phase I study, conducted between 2015-2020 across 25 countries, surveyed children and adolescents and found that the prevalence of asthma symptoms was 9.1% in children and 11.0% in adolescents, with lower-income countries generally showing lower prevalence rates than higher-income countries[6]. The latest research based on GBD 2021 revealed that the global age-standardized asthma prevalence rate was 3,340 per 100,000 population in 2021, with higher prevalence among males under 20 years of age. While incidence and prevalence rates showed positive correlations with the socio-demographic index, mortality and disability-adjusted life years demonstrated negative correlations[7]. However, with the increasing availability of global asthma health data and updates to the GBD database, a comprehensive analysis of the global asthma burden is warranted[8].

Therefore, this study aims to systematically analyze the evolving patterns of the global disease burden of asthma among children and adolescents (aged 0-19 years) from 1990 to 2023, utilizing the latest Global Burden of Disease Study database. This study will comprehensively reveal the complex landscape of global childhood and adolescent asthma prevention and control. The findings will provide crucial scientific evidence for developing age- and sex-specific asthma prevention and control strategies, optimizing resource allocation, and promoting health equity, with significant implications for achieving the United Nations Sustainable Development Goals related to child health.

## 2 Methods

### 2.1 Data Sources and Indicator Calculations

Data for this study were obtained from the Global Burden of Disease (GBD) Study. We downloaded epidemiological data for childhood and adolescent asthma (ages 0-19) from 1990 to 2023 for global and 21 regions through the GBD website, including crude incidence rates, crude prevalence rates, age-standardized incidence rates (ASIR), age-standardized prevalence rates (ASPR), age-standardized mortality rates (ASMR), and age-standardized disability-adjusted life years (DALYs) rates. Corresponding demographic data were also downloaded for standardization calculations. Key indicators included: crude prevalence rate, crude incidence rate, ASPR, ASIR, ASMR, and age-standardized DALYs rate (ASDR).

### 2.2 Descriptive Analysis of Disease Burden

Descriptive analyses were conducted for childhood and adolescent asthma incidence, prevalence, mortality, and DALYs rates globally and across 21 regions from 1990 to 2023. Prevalent cases, incident cases, deaths, DALYs, crude rates, age-standardized rates, and percentage changes from 1990 to 2023 were calculated. Disease burden maps were generated to visualize the global distribution patterns.

### 2.3 Joinpoint Regression Analysis

Joinpoint regression models were applied to analyze temporal trends in age-standardized rates from 1990-2023. Standard errors were calculated using R software, data were imported into Joinpoint software with time as independent variable, age-standardized rates as dependent variable, region as grouping variable, maximum 5 joinpoints, 95% confidence intervals, and parametric testing. Results were expressed as annual percentage change (APC).

### 2.4 Age-Period-Cohort Analysis

To explore temporal trends, we excluded 1990-1991 data and divided 1992-2023 data into 5-year periods for APC analysis. We calculated effects of age, period, and cohort on asthma incidence, created relative risk plots, and calculated local drift percentages.

### 2.5 Decomposition Analysis

Das Gupta decomposition method was applied to analyze contributions of population, epidemiological, and age structure factors to prevalence changes globally and by GBD/SDI regions. Frontier Analysis: Data envelopment analysis evaluated relationships between disease burden and sociodemographic index (SDI). FDH models fitted nonlinear production frontiers, LOWESS generated smooth frontiers, and super-efficiency points were excluded to avoid outlier effects.

### 2.6 Health Inequality Analysis

Slope index of inequality (SII) and concentration index (CI) were calculated for age-standardized DALYs rates, with SII reflecting absolute inequality and CI relative inequality. We calculated and compared 1990 and 2023 values.

### 2.7 Prediction Analysis

The Bayesian Age-Period-Cohort (BAPC) model was selected for time series analysis and prediction. Based on the established BAPC model, predictions were made for age-standardized rates and sex groups of childhood and adolescent asthma from 2024-2038.

### 2.8 Statistical Analysis

All analyses were performed using R 4.2.0, with BAPC package for Bayesian models. *P*<0.05 was considered statistically significant.

## 3 Results

### 3.1 Temporal and Regional Trends in Childhood and Adolescent Asthma Disease Burden

In 2023, the global prevalence of asthma among children and adolescents reached 130.52 million cases (95% UI: 89.518-190.284 million), with an age-standardized prevalence rate of 1,789.9 per 100,000 population (95% UI: 1,226.4-2,612.1). In the same year, asthma-related disability-adjusted life years (DALYs) totaled 6.5496 million (95% UI: 4.247-9.925 million), corresponding to an age-standardized DALYs rate of 90.5 per 100,000 (95% UI: 58.7-136.9). The global incident cases numbered 43.827 million (95% UI: 25.883-69.250 million), with an age-standardized incidence rate of 622.2 per 100,000 (95% UI: 369.2-981.0). Global deaths attributed to asthma were 16,300 (95% UI: 11,200-23,600), with an age-standardized mortality rate of 0.230 per 100,000 (95% UI: 0.158-0.333) (Table 1-4).

**Table 1.**
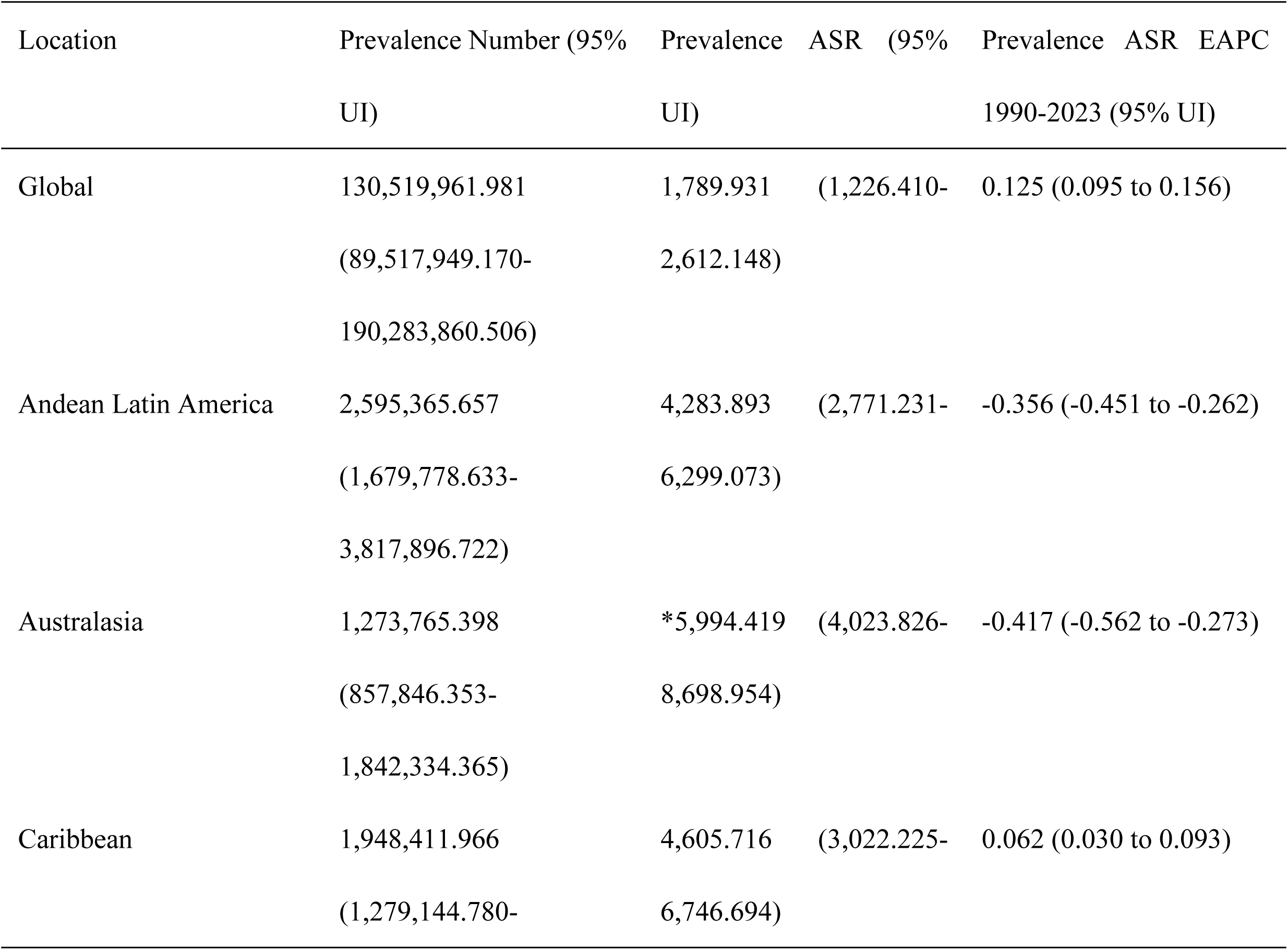
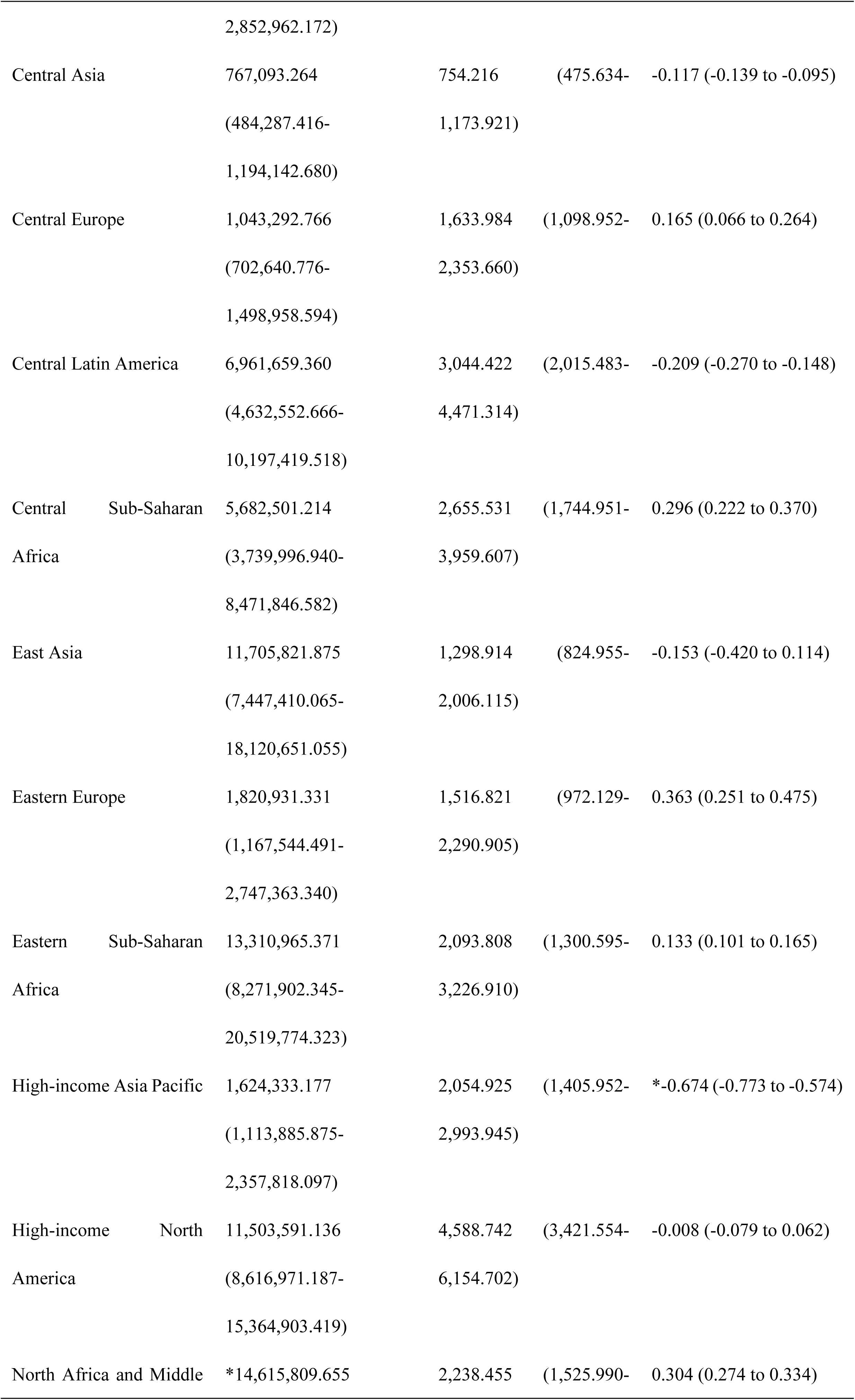
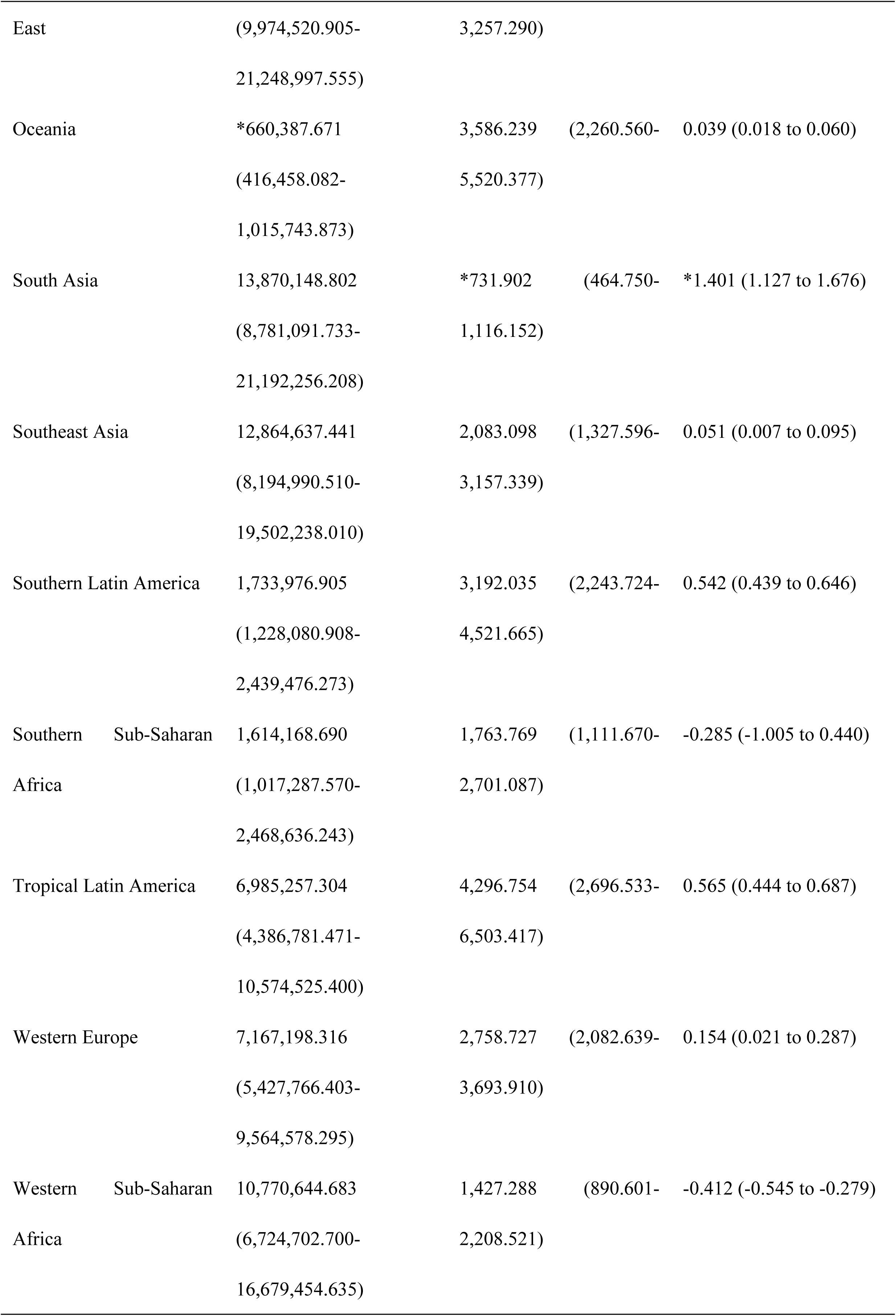
Prevalent cases and ASPR of childhood and adolescent asthma globally and in 21 GBD regions in 2023, with EAPC from 1990-2023.

**Table 2.**
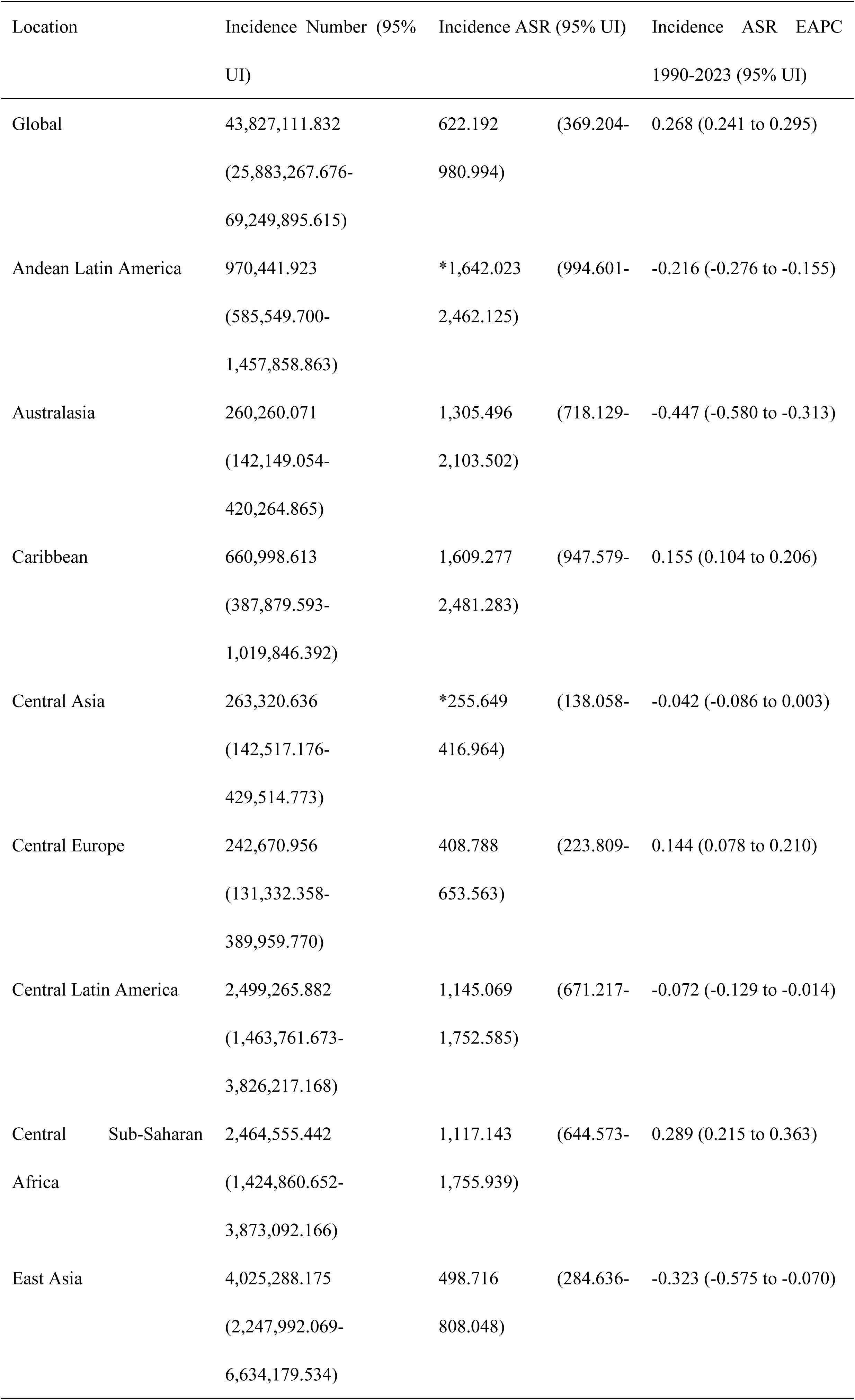
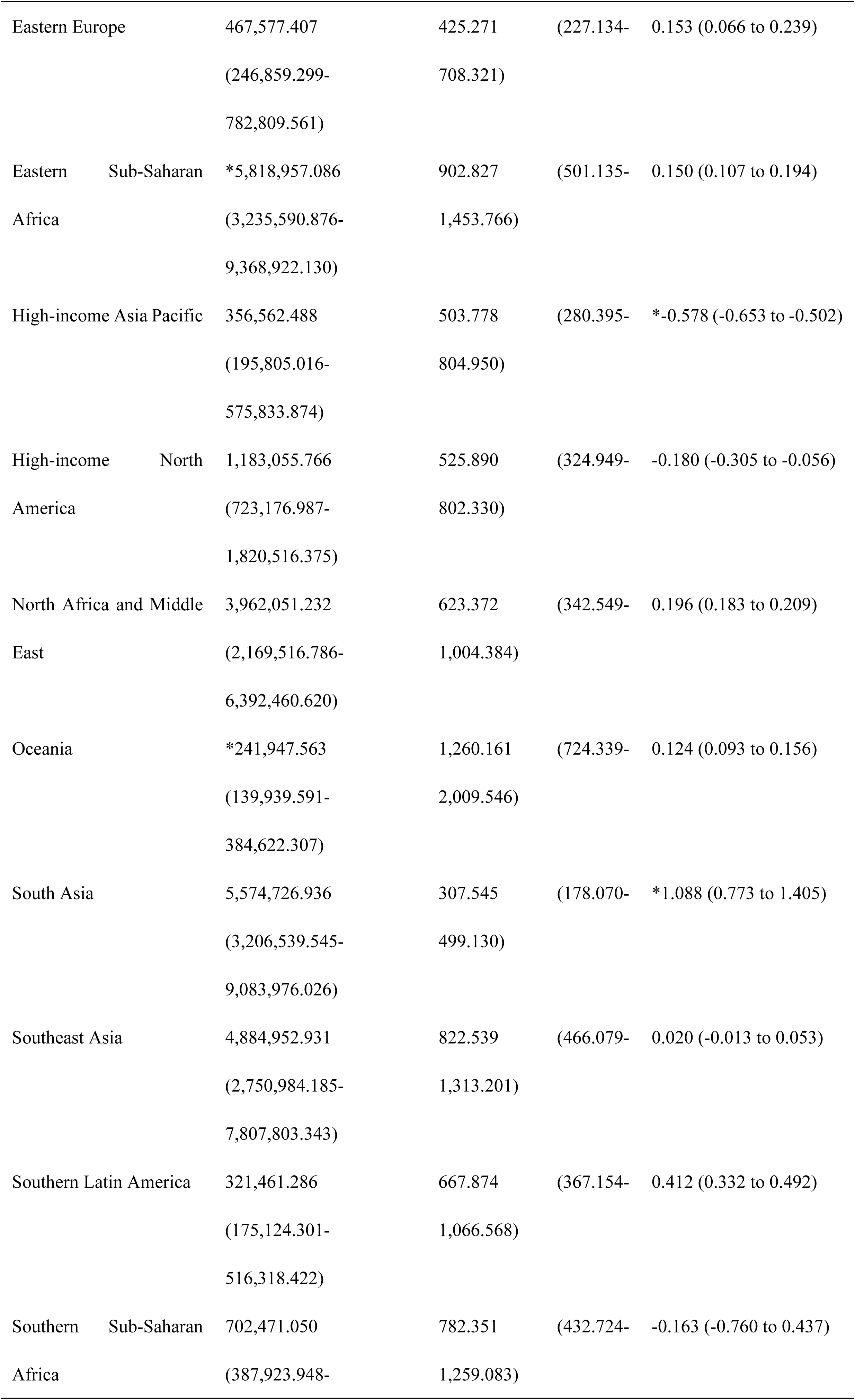
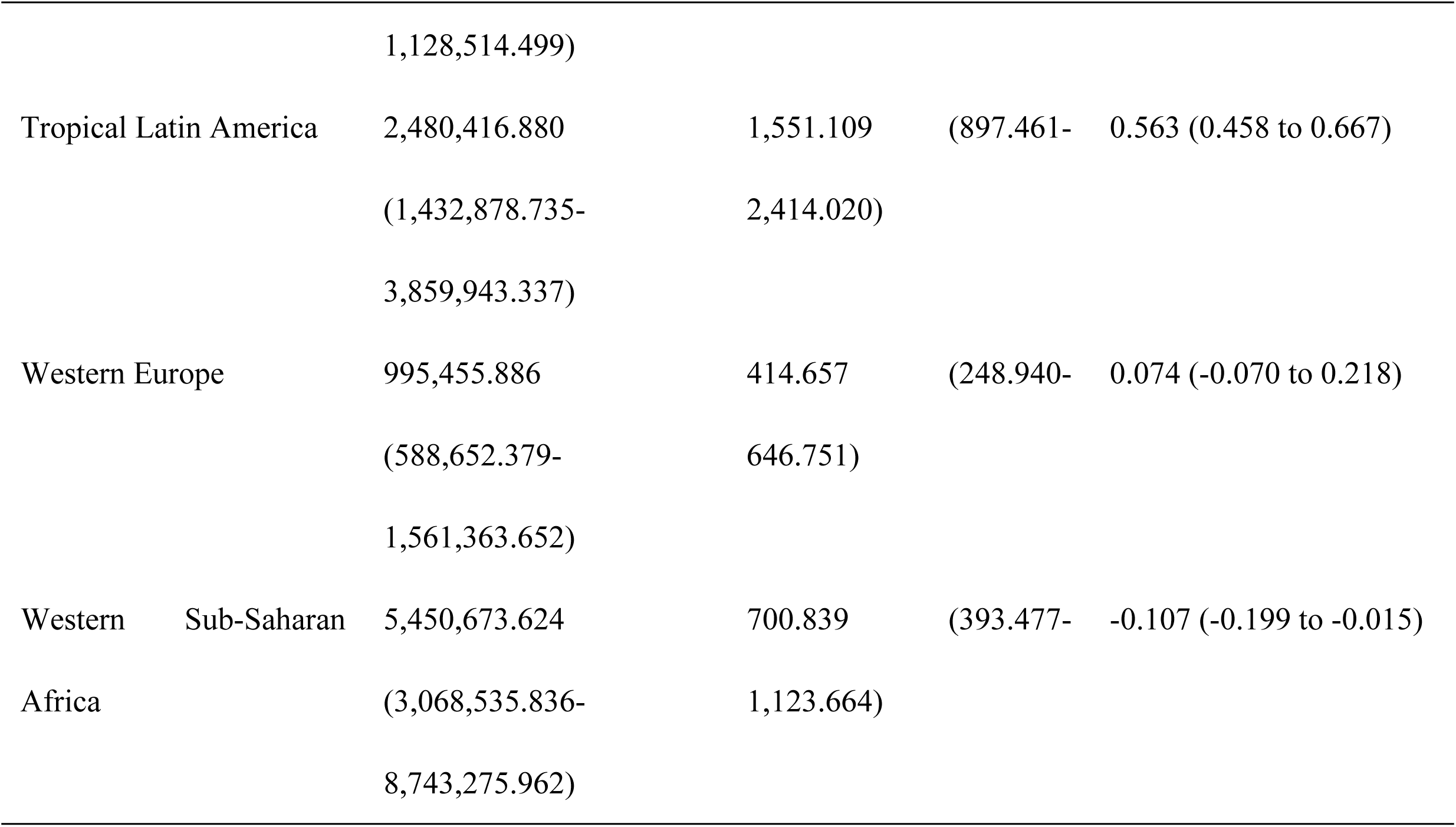
Incident cases and ASIR of childhood and adolescent asthma globally and in 21 GBD regions in 2023, with EAPC from 1990-2023.

**Table 3.** Deaths and ASMR of childhood and adolescent asthma globally and in 21 GBD regions in 2023, with EAPC from 1990-2023.

| Location | Deaths Number (95% UI) | Deaths ASR (95% UI) | Deaths ASR EAPC 1990-2023 (95% UI) |
| --- | --- | --- | --- |
| Global | 16,269.813 (11,200.543-23,555.737) | 0.230 (0.158-0.333) | -1.578 (-1.671 to -1.486) |
| Andean Latin America | 68.743 (41.840-108.530) | 0.115 (0.070-0.183) | -6.671 (-7.245 to -6.093) |
| Australasia | 18.037 (15.480-21.142) | 0.081 (0.070-0.095) | -3.292 (-3.603 to -2.981) |
| Caribbean | 145.304 (91.107-220.915) | 0.350 (0.219-0.533) | -1.749 (-1.949 to -1.549) |
| Central Asia | 53.141 (40.889-68.718) | 0.053 (0.041-0.068) | -2.194 (-2.617 to -1.768) |
| Central Europe | *4.197 (3.442-5.195) | *0.006 (0.005-0.007) | -6.333 (-6.577 to -6.088) |
| Central Latin America | 189.262 (149.197-237.594) | 0.088 (0.069-0.111) | -5.650 (-5.905 to -5.395) |
| Central Sub-Saharan Africa | 866.021 (533.185-1,373.576) | 0.406 (0.250-0.645) | -1.525 (-1.624 to -1.427) |
| East Asia | 140.597 (81.965-227.041) | 0.015 (0.009-0.024) | -5.887 (-6.194 to -5.578) |
| Eastern Europe | 17.964 (14.506-22.381) | 0.014 (0.011-0.017) | -6.187 (-6.544 to -5.828) |
| Eastern Sub-Saharan Africa | 2,691.154 (1,639.380-4,227.207) | 0.419 (0.255-0.658) | -1.545 (-1.638 to -1.452) |
| High-income Asia Pacific | 12.059 (7.727-23.197) | 0.015 (0.009-0.028) | *-9.564 (-9.833 to -9.294) |
| High-income North America | 212.739 (179.345-254.432) | 0.083 (0.070-0.100) | -1.608 (-1.837 to -1.378) |
| North Africa and Middle East | 775.979 (459.121-1,255.649) | 0.121 (0.071-0.196) | -4.376 (-4.510 to -4.242) |
| Oceania | 55.090 (29.363-93.127) | 0.311 (0.166-0.525) | -1.295 (-1.424 to -1.166) |
| South Asia | 3,301.267 (2,066.143-4,909.908) | 0.169 (0.105-0.252) | -2.480 (-2.604 to -2.355) |
| Southeast Asia | 3,165.927 (2,036.053-4,693.723) | *0.521 (0.333-0.776) | -1.012 (-1.073 to -0.951) |
| Southern Latin America | 10.035 (7.681-12.941) | 0.017 (0.013-0.022) | -4.843 (-5.157 to -4.528) |
| Southern Sub-Saharan Africa | 421.539 (279.781-625.301) | 0.484 (0.320-0.719) | *-0.825 (-1.010 to -0.639) |
| Tropical Latin America | 144.959 (120.258-171.055) | 0.089 (0.074-0.105) | -4.997 (-5.409 to -4.584) |
| Western Europe | 71.961 (63.249-82.399) | 0.027 (0.024-0.031) | -4.070 (-4.230 to -3.911) |
| Western Sub-Saharan Africa | *3,903.839 (2,432.459-6,121.392) | 0.489 (0.305-0.766) | -1.789 (-2.092 to -1.485) |

**Table 4.**
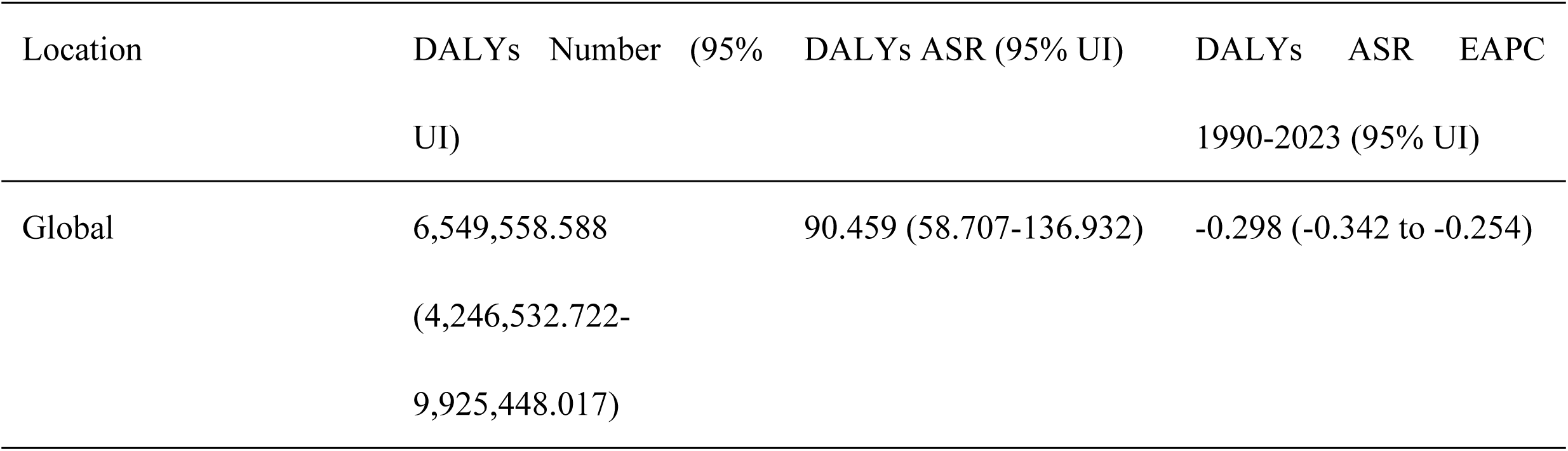
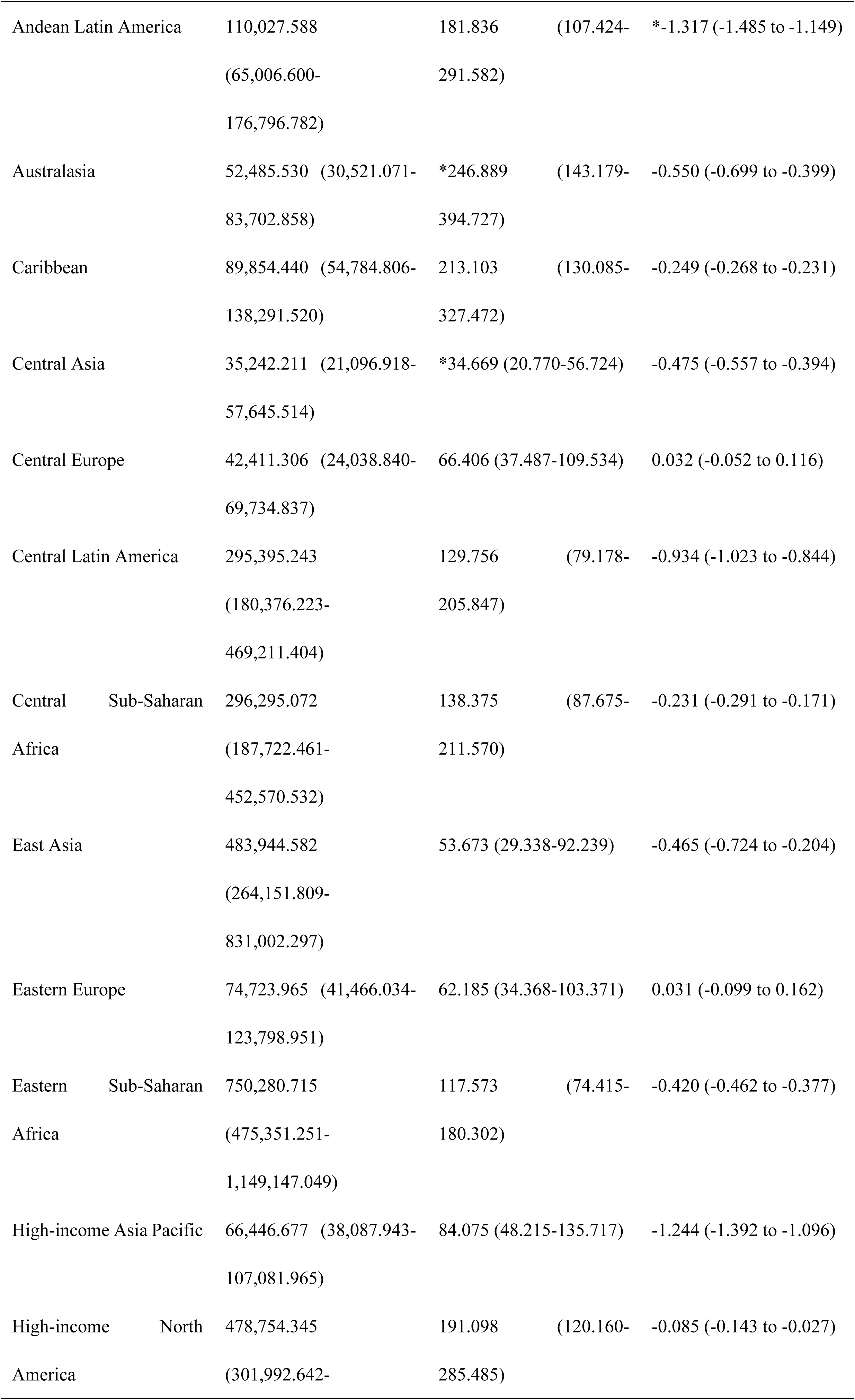
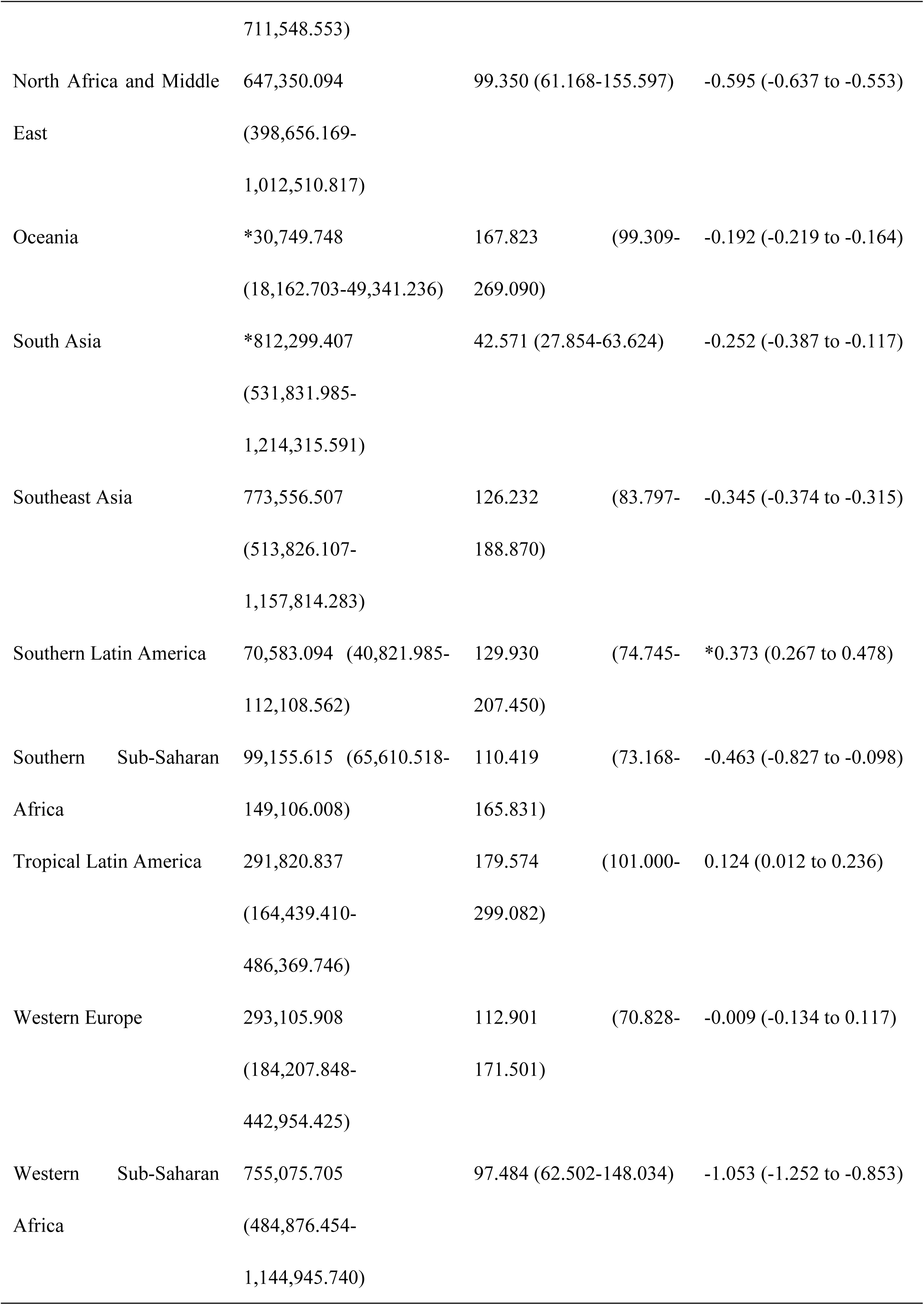
DALYs and ASDR of childhood and adolescent asthma globally and in 21 GBD regions in 2023, with EAPC from 1990-2023.

Among the 21 GBD regions, North Africa and the Middle East exhibited the highest prevalence in 2023, with 14.616 million cases, followed by South Asia (13.870 million) and Eastern Sub-Saharan Africa (13.311 million). Australasia demonstrated the highest age-standardized prevalence rate (5,994.4 per 100,000), followed by the Caribbean (4,605.7 per 100,000), High-income North America (4,588.7 per 100,000), and Tropical Latin America (4,296.8 per 100,000). South Asia recorded the lowest age-standardized prevalence rate at 731.9 per 100,000 (Figure 1A). In 2023, Eastern Sub-Saharan Africa reported the highest number of incident cases (5.819 million), followed by South Asia (5.575 million). The highest age-standardized incidence rates were observed in Andean Latin America (1,642.0 per 100,000), the Caribbean (1,609.3 per 100,000), and Tropical Latin America (1,551.1 per 100,000), while Central Asia recorded the lowest rate (255.6 per 100,000) (Figure 1B). Australasia exhibited the highest age-standardized DALYs rate (246.9 per 100,000), followed by the Caribbean (213.1 per 100,000), High-income North America (191.1 per 100,000), and Andean Latin America (181.8 per 100,000) (Figure 1D). The highest age-standardized mortality rates were observed in Southeast Asia (0.521 per 100,000) and Western Sub-Saharan Africa (0.489 per 100,000), while Central Europe recorded the lowest rate (0.006 per 100,000) (Figure 1C).

**Figure 1.**
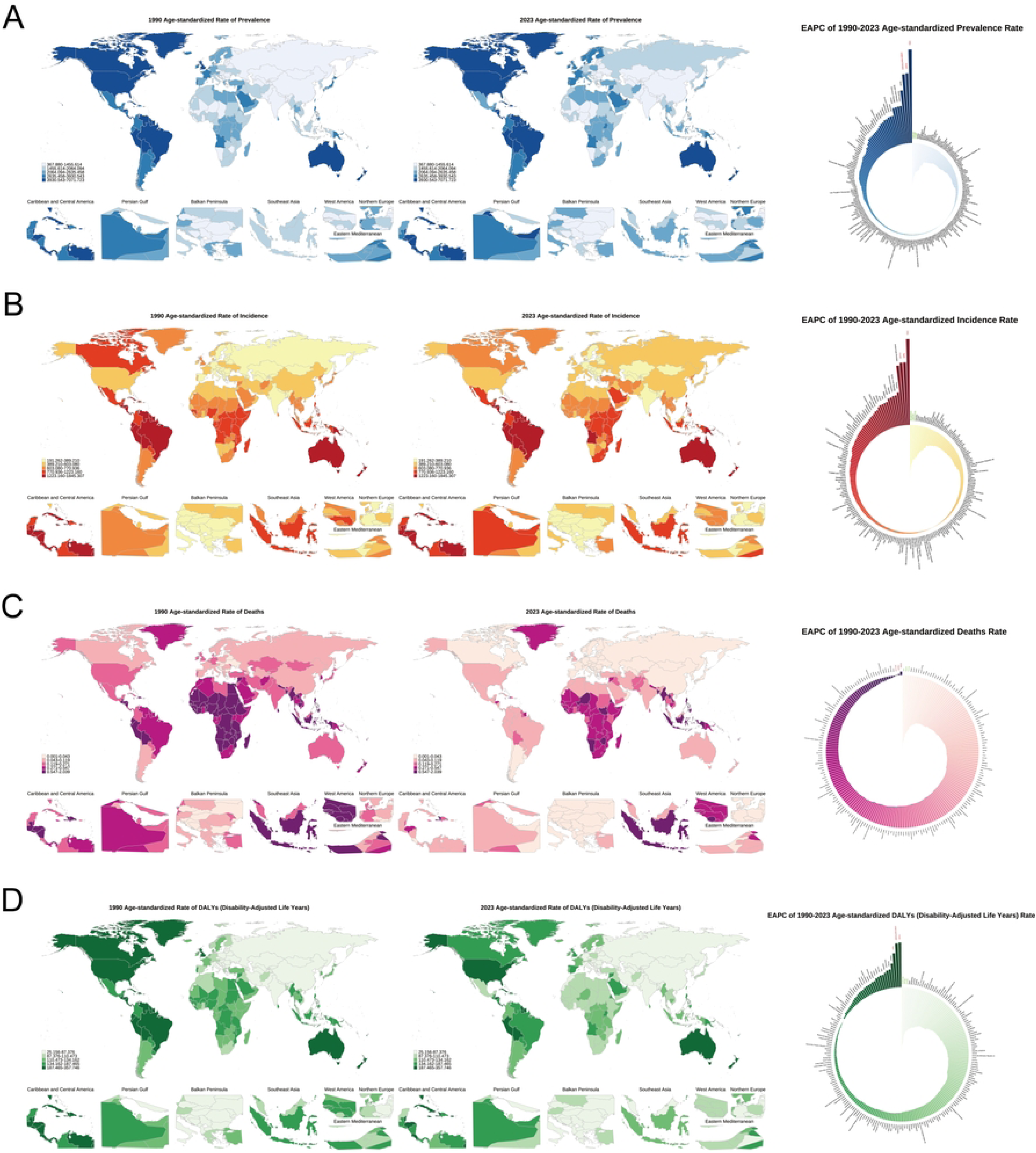
Spatial distribution and temporal trends: Spatial distribution and temporal trends of the global disease burden of childhood and adolescent asthma (1990-2023). Distribution and changes in global age-standardized prevalence (A), incidence (B), mortality (C), and DALYs rates (D) of childhood and adolescent asthma (per 100,000 population): (Left) Distribution in 1990; (Middle) Distribution in 2023; (Right) Estimated annual percentage change (EAPC) from 1990 to 2023. Color intensity indicates the level of disease burden. In the EAPC maps, red indicates the top three countries with the fastest increase, and green indicates the three countries with the fastest decrease.

From 1990 to 2023, the global age-standardized prevalence rate of asthma showed a gradual upward trend, with an estimated annual percentage change (EAPC) of 0.125% (95% UI: 0.095-0.156). South Asia experienced the most significant increase (EAPC: 1.401%), while High-income Asia Pacific showed the most pronounced decline (EAPC: -0.674%). The global age-standardized incidence rate increased during the same period, with an EAPC of 0.268% (95% UI: 0.241-0.295). South Asia demonstrated the most substantial increase in incidence (EAPC: 1.088%), whereas High-income Asia Pacific showed the steepest decline (EAPC: -0.578%). Conversely, the global age-standardized

DALYs rate exhibited a downward trend, with an EAPC of -0.298% (95% UI: -0.342 to -0.254). Southern Latin America showed the most significant increase (EAPC: 0.373%), while Andean Latin America experienced the most substantial decrease (EAPC: -1.317%). Notably, the global asthma mortality rate demonstrated a significant downward trend, with an EAPC of -1.578% (95% UI: - 1.671 to -1.486). All 21 regions showed declining mortality trends, with High-income Asia Pacific achieving the most remarkable improvement (EAPC: -9.564%).

### 3.2 Age and Sex Patterns in Childhood and Adolescent Asthma Disease Burden

Comprehensive analysis revealed significant age and sex differential patterns. Males showed higher prevalence and disease burden across age groups, with sex reversal trends emerging during adolescence. Prevalent cases peaked at ages 5-9 for males (22.89 million) and 10-14 for females (17.16 million); prevalence rates peaked at ages 5-9 for males (6,349.1/100,000) and 10-14 for females (5,187.0/100,000), with females first exceeding males at ages 15-19 (4,591.1 vs 4,173.7/100,000) (Figure 2A-B). Incidence declined significantly with age, highest in children <5 years (males 9.43 million, females 7.42 million cases; rates 2,849.0 and 2,382.8/100,000 respectively), lowest at ages 15-19 with females (729.0/100,000) exceeding males (617.2/100,000) (Figure 2C-D). Mortality showed U-shaped distribution, higher at <5 and 15-19 years; males slightly higher at <5 years (1.039 vs 0.994/100,000), lowest at 5-9 years, rising again at 15-19 with females (0.709/100,000) exceeding males (0.635/100,000) (Figure 2E-F). DALYs burden was heaviest at ages 5-9 for males (1.03 million person-years) and 10-14 for females (0.79 million); DALYs rates showed similar patterns, peaking at 5-9 for males (285.0/100,000) and 10-14 for females (240.0/100,000), with notable sex reversal at 15-19 where females (233.5/100,000) exceeded males (213.3/100,000) (Figure 2G-H), indicating important sex transition phenomena with implications for age- and sex-specific control strategies.

**Figure 2.**
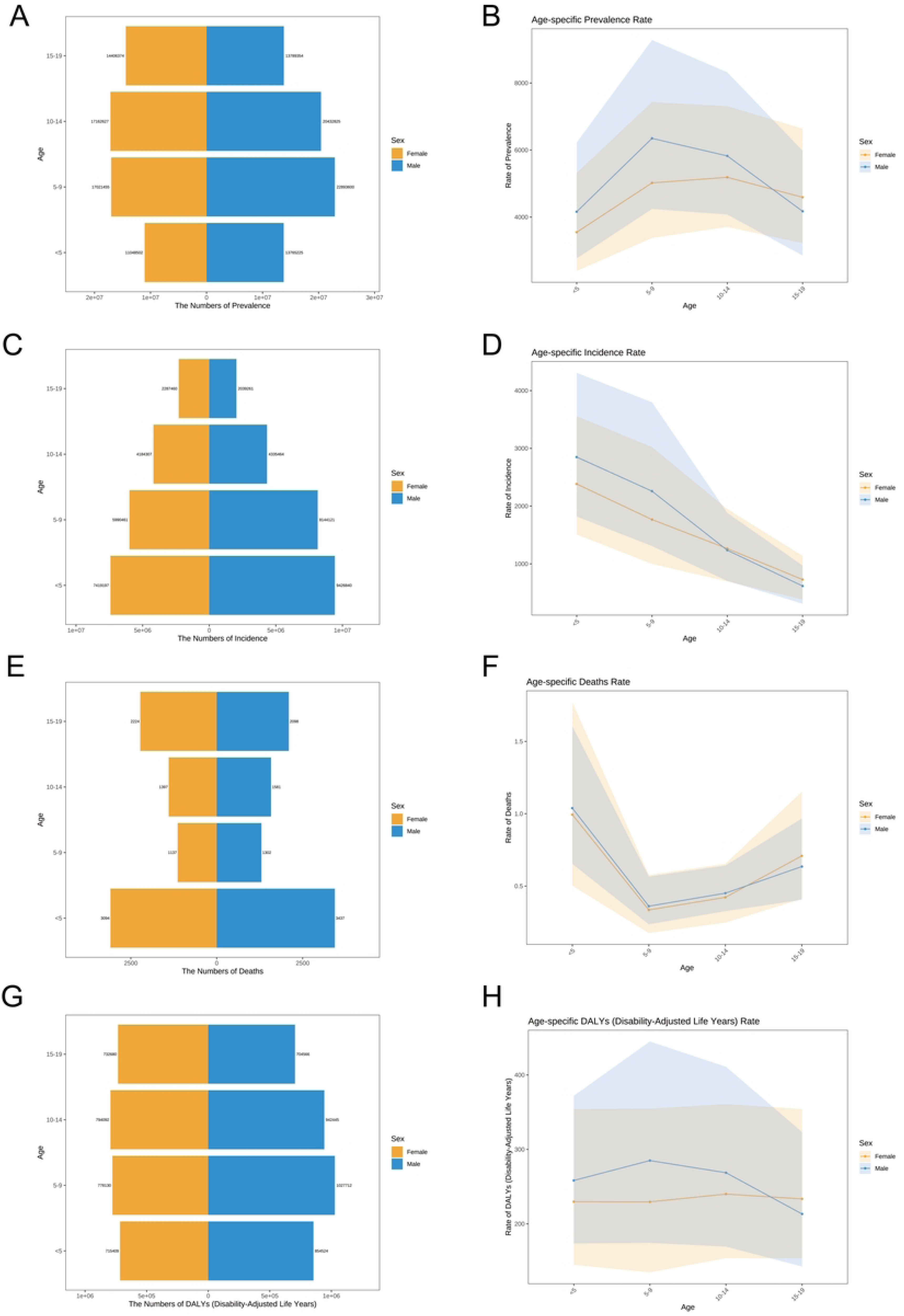
Age-specific distribution characteristics of global childhood and adolescent asthma disease burden. A-B: Age-specific prevalence rates and distribution. C-D: Age-specific incidence rates and distribution. E-F: Age-specific mortality rates and distribution. G-H: Age-specific disability-adjusted life years (DALYs) rates and distribution.

### 3.3 Joinpoint Regression Analysis

Joinpoint regression analysis of long-term trends from 1990-2023 showed incidence increased from 582.91 to 622.19 per 100,000 (6.74% increase), prevalence from 1,745.71 to 1,789.93 (2.53% increase), indicating continued growth in affected populations (Figure 3A-B). Conversely, mortality decreased dramatically from 0.441 to 0.230 per 100,000 (47.85% decline), DALYs rate from 99.78 to 90.46 per 100,000 (9.34% decline), reflecting significant improvements in management and treatment (Figure 3C-D). Analysis identified 2020-2021 as a critical turning point, with relatively stable trends before but reversals during this period: prevalence and incidence peaked in 2020 then declined, while mortality and DALYs rates rebounded 5.50% and 0.62% after reaching historic lows, possibly related to COVID-19 impacts. Sex analysis showed males consistently higher than females but gaps narrowing, particularly mortality sex ratio declining from 1.69 in 1990 to 1.01 in 2023, essentially eliminating sex differences. These findings indicate that while lethality and disease burden have markedly decreased, continued rising incidence and recent indicator rebounds suggest need for strengthened prevention and control strategies.

**Figure 3.**
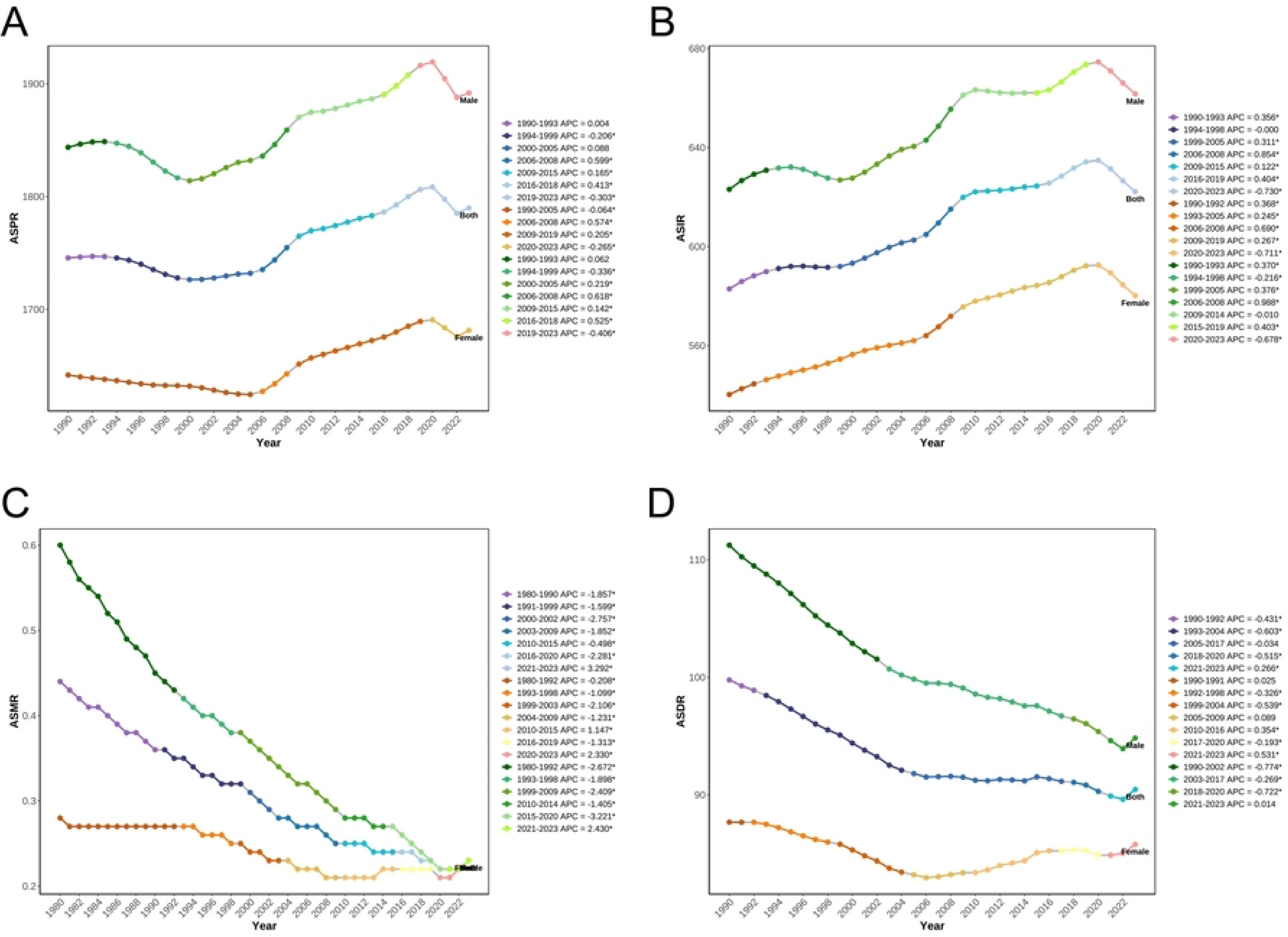
Joinpoint regression analysis of global childhood and adolescent asthma 1990-2023. (A) ASPR changes 1990-2023; (B) ASIR changes 1990-2023; (C) ASMR changes 1990-2023; (D) ASDR changes 1990-2023.

### 3.4 Age-Period-Cohort Effect Analysis

Comprehensive APC analysis in ages 0-19 revealed differential epidemiological evolution. Age effects showed inverted U-shaped prevalence peaking at 7.5 years (5,699.3/100,000), incidence declining from 2,528.4/100,000 at 2.5 years to 597.6/100,000 at 17.5 years (76.4% decrease), U-shaped mortality higher in early childhood (1.345/100,000) and adolescence (0.625/100,000), and inverted V-shaped DALYs peaking at 7.5 years (275.21/100,000 person-years). Period effects showed clear divergence: with 2006 as reference, prevalence and incidence increased 4.1% and 7.4% by 2021, while mortality decreased 33.1% from 1996-2021 (RR from 1.266 to 0.847), DALYs relatively stable (8.7% fluctuation). Cohort effects showed bidirectional trends: post-2009 birth cohorts had 5.5% increased prevalence and incidence risk, while mortality and DALYs improved continuously, with 2019 cohort showing 28.1% and 10% decreases versus 1999. Local drift analysis showed positive growth for prevalence and incidence across age groups (annual 0.186%-0.331%), negative improvement for mortality and DALYs (mortality -1.524% annually, DALYs -0.429% annually in younger groups) (Figure 4A-D). This “rising incidence but improving prognosis” separation reflects dual challenges: environmental deterioration and lifestyle changes drive continued risk increases, particularly in newer generations, while standardized care and medical advances significantly reduce disease severity and mortality risk, emphasizing urgency of strengthening primary prevention alongside clinical management.

**Figure 4.**
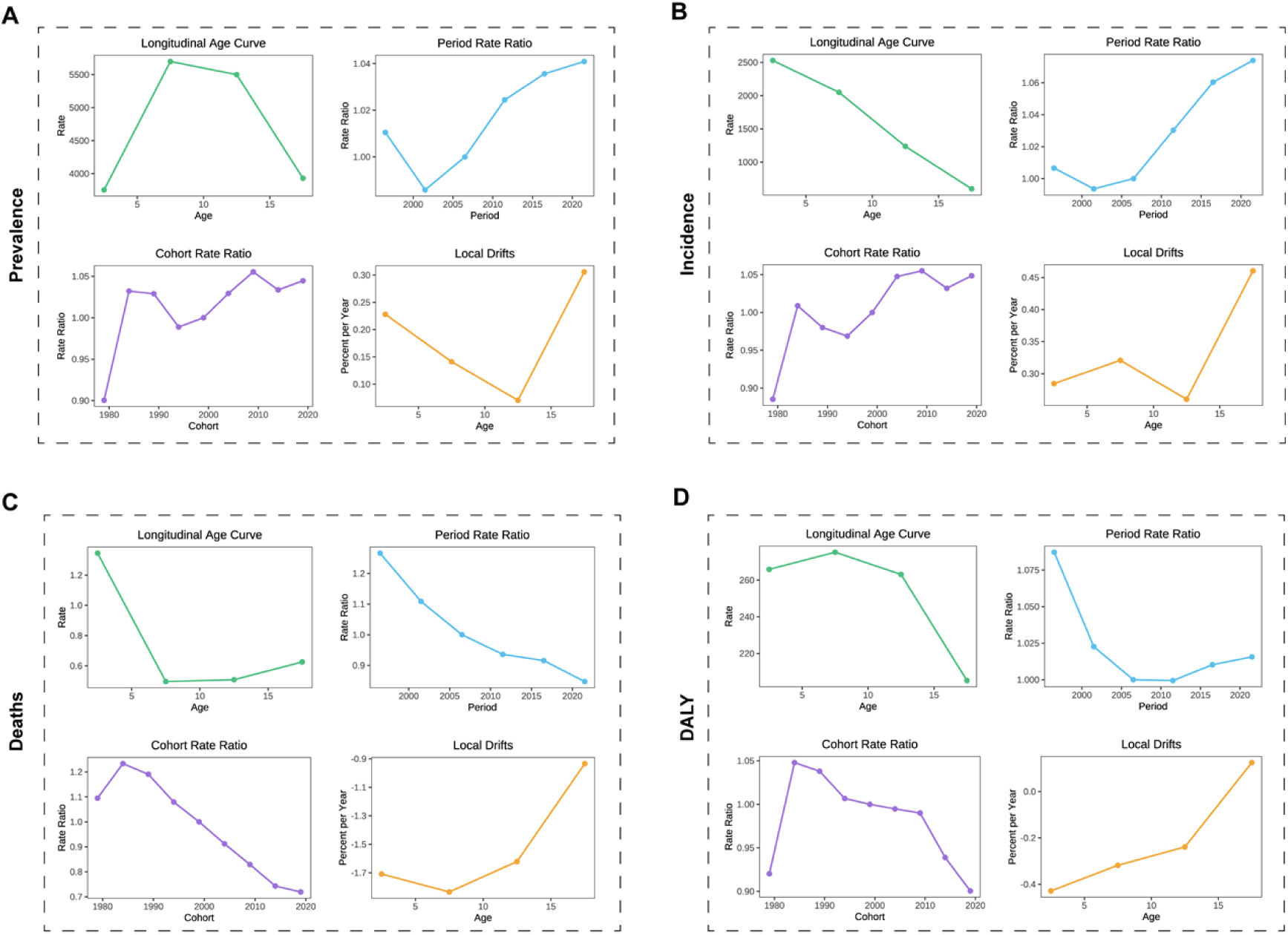
Age-period-cohort effect analysis of global childhood and adolescent asthma disease burden measures 1990-2023. (A-D) Age, period, cohort effects and local drift analysis results for prevalence, incidence, mortality, and DALYs.

### 3.5 Decomposition Analysis

Decomposition analysis revealed that incidence and prevalence increases were primarily driven by population growth, contributing 82.1% and 83.5% respectively, with epidemiological changes secondary (32.5% and 12.9%), while population aging negatively impacted incidence (-14.6%) with weak effect on prevalence (3.6%) (Figure 5A-B). Globally, epidemiology has a negative impact on mortality (contributing a 144.9% decrease), although population growth brings a 54.8% positive pressure; The DALYs rate was also driven by epidemiological improvement (contributing a 136.0% decrease), but was significantly offset by population growth (contributing a 241.2% increase) (Figure 5C-D). This comprehensive analysis reveals dual challenges: medical advances have significantly reduced lethality and severity, while population growth continues driving absolute case increases. Future control strategies must maintain and enhance treatment quality while emphasizing prevention measures, particularly addressing population growth-related burden increases.

**Figure 5.**
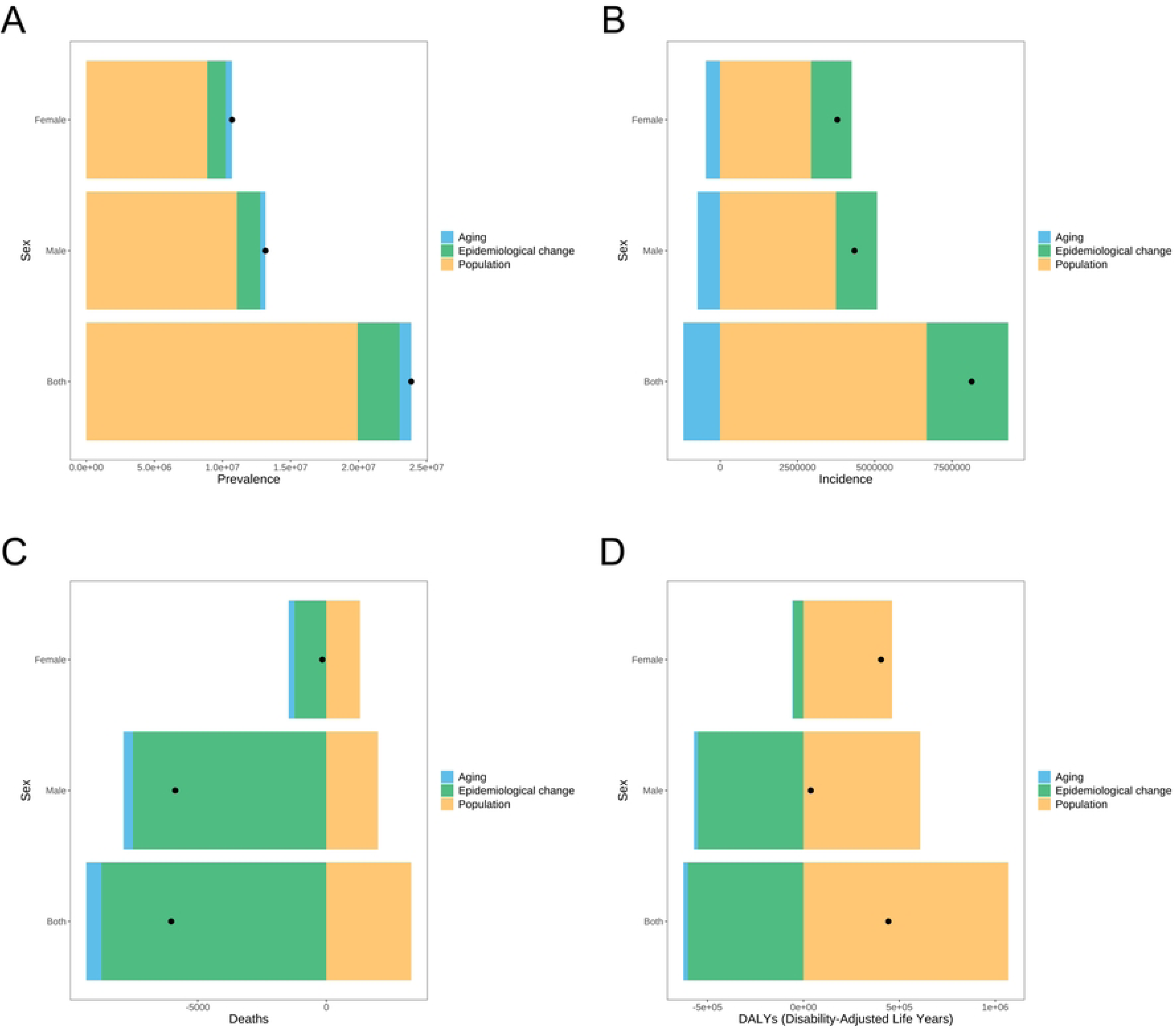
Decomposition analysis of changes in childhood and adolescent asthma prevalence (A), incidence (B), mortality (C), and DALYs rates (D) burden 1990-2023.

### 3.6 Relationship Between Disease Burden and Sociodemographic Development

Figure 6A displays the relationship between age-standardized DALYs rates and Human Development Index (HDI) globally. The first panel presents time series data from 1990-2020, showing evolution across four time points (light blue 1990 to dark blue 2020). Most notable is the black curve connecting countries with highest asthma burden at each HDI level, forming an “efficiency frontier” upper bound. This curve remains relatively low at low-medium HDI (0.3-0.5), gradually rising with increasing HDI, reaching highest levels at high HDI (>0.8), showing positive correlation.

**Figure 6.**
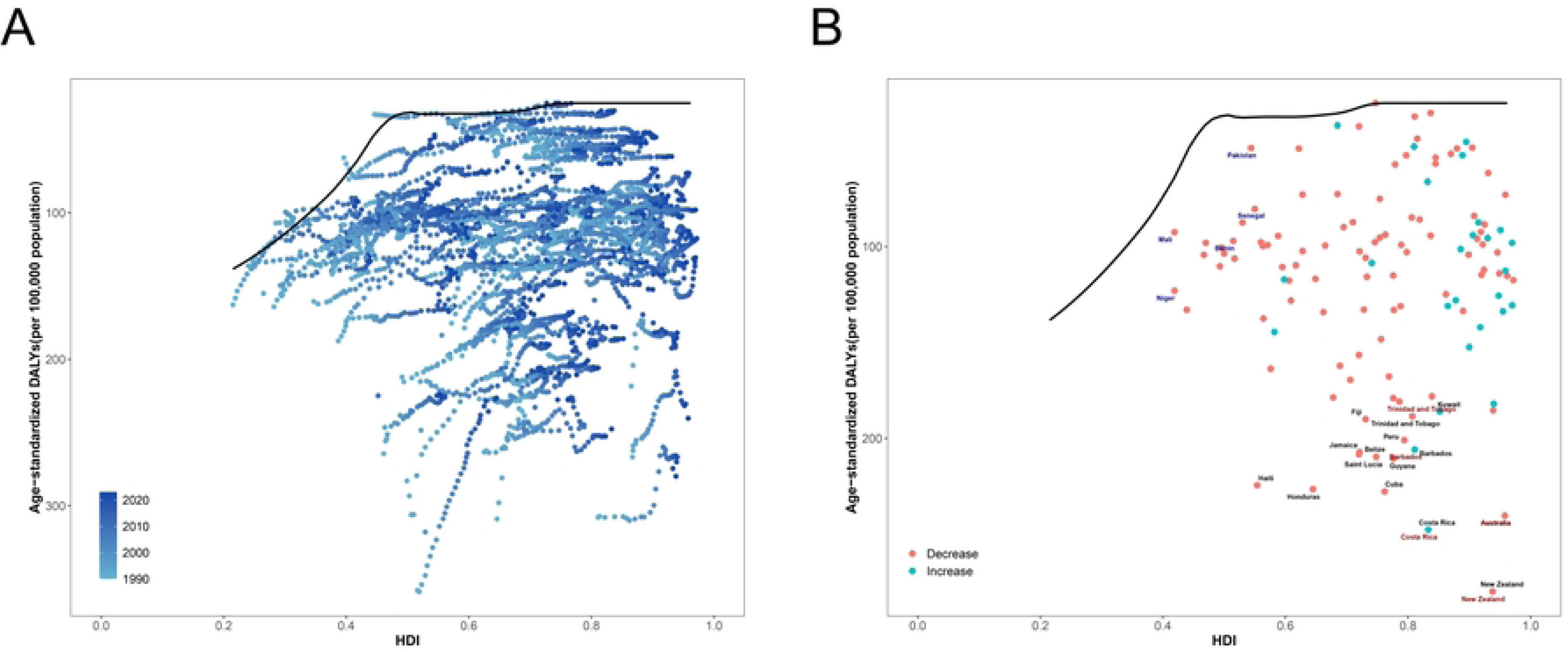
Relationship between disease burden and sociodemographic development: Relationship between SDI and age-standardized DALYs rate of childhood and adolescent asthma 1990-2023. (A) Scatter plot of SDI vs age-standardized DALYs rate for 124 countries 1990-2023. Black curve represents frontier production function fit, indicating minimum DALYs rate at given HDI level. Different colored points represent different years. (B) Trends in age-standardized DALYs rates for selected countries in 2023. Black solid line is frontier production function fit, colored dotted lines show individual country DALYs rate changes. Black text indicates 15 points with largest distance differences; blue text indicates 5 low-HDI countries with smallest distance differences; red text indicates 5 high-HDI countries with largest distance differences.

Figure 6B details specific country performance and trends. Red-marked countries (New Zealand 280.0, Australia 240.3, Costa Rica 247.6, etc.) represent regions with declining but still high asthma burden. Cyan-marked countries (Pakistan 48.5, Mali 92.4, Senegal 87.5, Niger 123.0, Benin 97.0) represent increasing burden regions. Notably, developed countries like New Zealand and Australia, despite very high HDI (>0.93), have the world’s highest childhood asthma DALYs burden, possibly related to high asthma prevalence, allergic constitution prevalence, and environmental factors. In contrast, high-HDI countries in Eastern Europe show relatively low burden (Lithuania 45.4, Estonia 48.4), indicating clear regional differences. This pattern challenges conventional wisdom, suggesting complex nonlinear relationships between socioeconomic development and asthma burden, with highly developed countries potentially facing more severe disease burden.

### 3.7 HDI and Childhood Asthma Disease Burden Relationship

Correlation analysis showed ASPR significantly correlated with HDI (Spearman R=0.244, P=0.001), suggesting increased prevalence in high-HDI regions (Figure 7A). ASIR showed weak negative correlation with HDI reaching statistical significance (Spearman R=-0.359, *P*<0.001), suggesting declining new cases in developed countries (Figure 7B). ASMR showed negative correlation with HDI reaching statistical significance (Spearman R=-0.837, *P*<0.001), suggesting declining mortality in developed countries (Figure 7C). ASDR correlation with HDI was not significant (Spearman R=-0.083, P=0.286), though high-HDI countries showed higher disease burden (Figure 7D).

**Figure 7.**
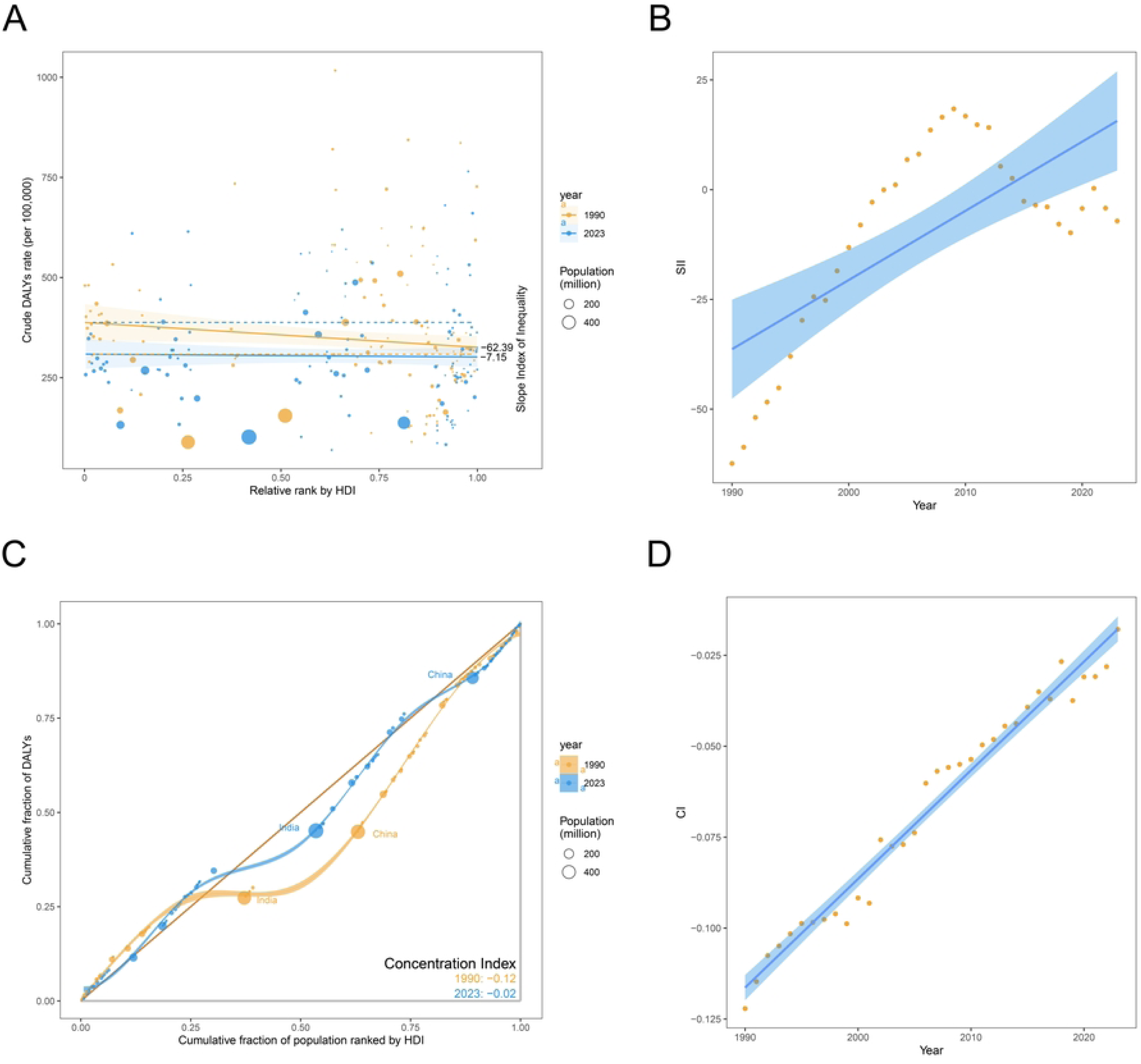
Relationship between HDI and childhood and adolescent asthma ASPR (A), ASIR (B), ASMR (C), and ASDR (D) 1990-2023.

### 3.8 Health Inequality Analysis

Health inequalities in childhood and adolescent asthma disease burden across countries with different Human Development Index (HDI) levels from 1990 to 2023 were analyzed using two core indicators: the Concentration Index (CI) and the Slope Index of Inequality (SII). It was found that the concentration index steadily increased from -0.12 in 1990 to -0.02 in 2023, indicating that the distribution of childhood and adolescent asthma-related DALYs across countries with different HDI levels has become increasingly equitable (Figure 8C-D). More remarkable improvements were observed in the Slope Index of Inequality, which dramatically decreased from -62.39 in 1990 to - 7.15 in 2023, representing a reduction of nearly 90% (Figure 8A). Time series analysis revealed that this improvement was primarily achieved during the period from the 1990s to the early 2000s, when the SII rapidly declined and approached zero at one point (Figure 8B). Subsequently, minor fluctuations were observed around 2010, but the overall level was maintained at a relatively low range. The negative values of both indicators suggest that countries with lower HDI still bear a relatively higher disease burden, although the degree of this inequality has been substantially diminished.

**Figure 8.**
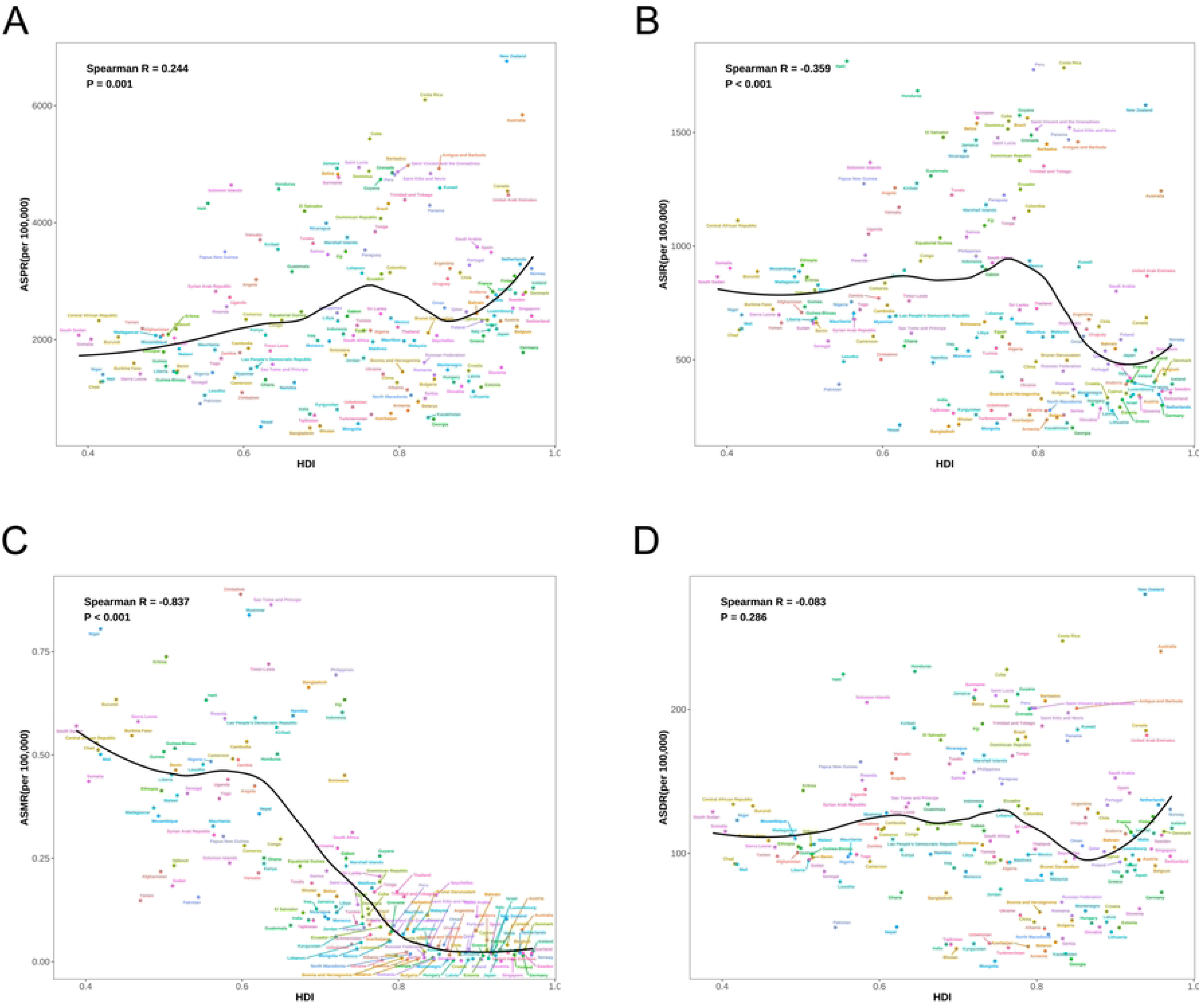
Health inequality analysis of age-standardized DALYs rate for childhood and adolescent asthma 1990-2023. (A) Trend in slope index of inequality for age-standardized DALYs rate 1990-2023. SII was -62.39 in 1990 and -7.15 in 2023; (B) Scatter plot of slope index of inequality for age-standardized DALYs rate 1990-2023. (C) Inequality curve for age-standardized DALYs rate. CI was -0.12 in 1990 and -0.02 in 2023; (D) Scatter plot of concentration index for age-standardized DALYs rate 1990-2023.

### 3.9 Future Projections of Childhood and Adolescent Asthma Disease Burden

To predict future global epidemiological patterns and disease burden trends of childhood and adolescent asthma, BAPC models were employed to project prevalence, incidence, mortality, and DALYs rates from 2024 to 2038. The age-standardized prevalence rate (ASPR) for childhood and adolescent asthma is projected to decrease from 1,785.7 per 100,000 population in 2023 to 1,751.8 per 100,000 in 2038. Significant sex differences were observed, with male prevalence projected to decline from 1,885.7 per 100,000 in 2023 to 1,829.7 per 100,000 in 2038, while female prevalence is expected to decrease from 1,679.9 per 100,000 in 2023 to 1,673.4 per 100,000 in 2038 (Figure 9A).

**Figure 9.**
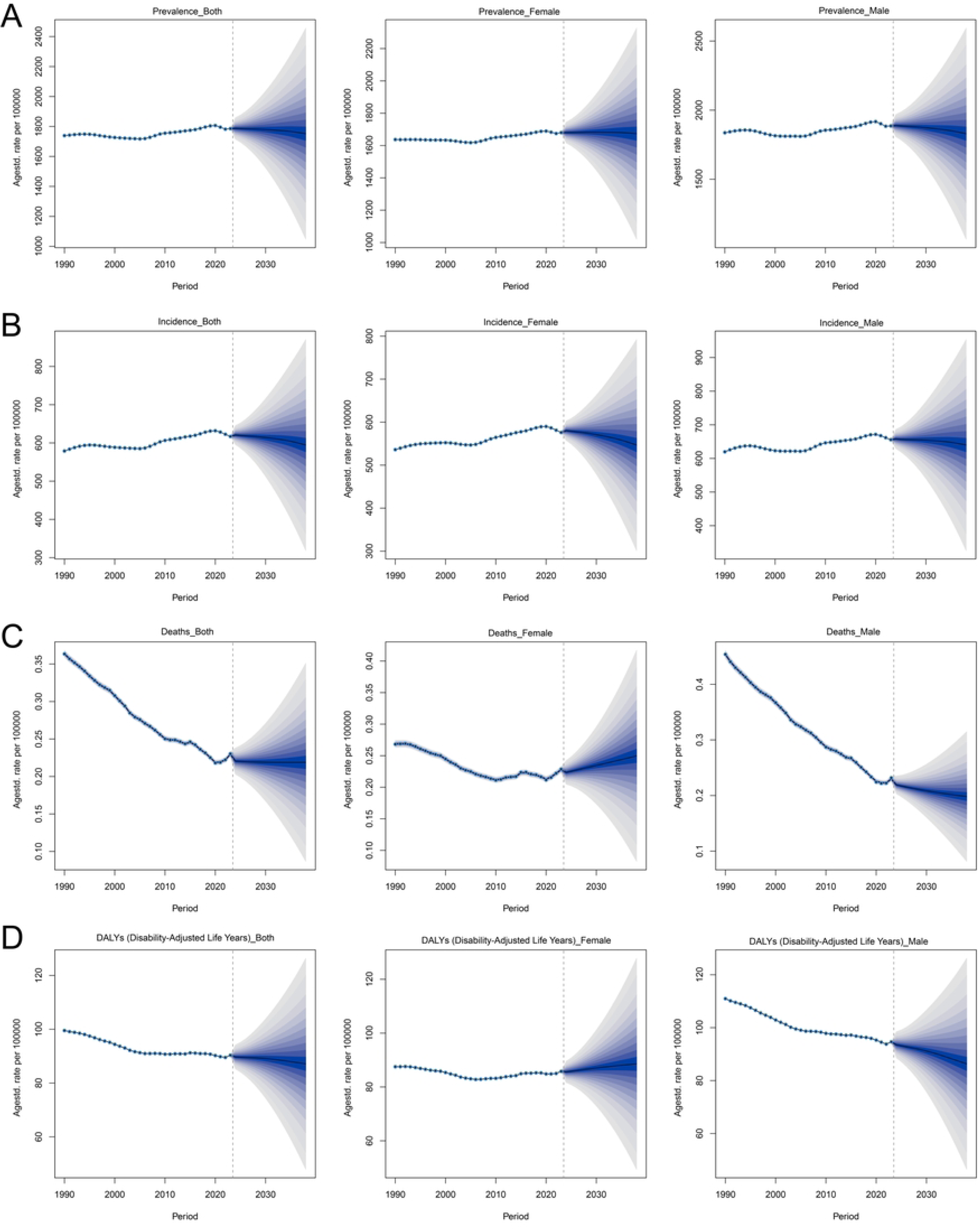
BAPC model-based projections of global childhood and adolescent asthma age-standardized prevalence (A), incidence (B), mortality (C), and DALYs (D) rates for 2024-2038.

Over the next 15 years, the age-standardized incidence rate (ASIR) for childhood and adolescent asthma is projected to show a declining trend, decreasing from 616.9 per 100,000 population in 2023 to 594.4 per 100,000 in 2038. Male incidence rates consistently remain higher than female rates, with male incidence projected to decline from 655.6 per 100,000 in 2023 to 639.8 per 100,000 in 2038, and female incidence from 575.9 per 100,000 to 546.8 per 100,000 (Figure 9B).

ASMR of childhood and adolescent asthma is projected to demonstrate a significant declining trend. The overall mortality rate is projected to decrease from 0.230 per 100,000 population in 2023 to 0.219 per 100,000 in 2038. Male mortality rates are expected to decline from 0.230 per 100,000 in 2023 to 0.198 per 100,000 in 2038, while female mortality rates are projected to increase from 0.228 per 100,000 in 2023 to 0.250 per 100,000 in 2038 (Figure 9C).

ASDR for childhood and adolescent asthma is projected to show an overall declining trend, decreasing from 90.3 per 100,000 population in 2023 to 87.1 per 100,000 in 2038. Significant sex differences were observed: male DALYs rates are projected to decline from 94.6 per 100,000 in 2023 to 86.3 per 100,000 in 2038, whereas female DALYs rates are expected to increase from 85.8 per 100,000 in 2023 to 88.6 per 100,000 in 2038 (Figure 9D).

## 4 Discussion

This study analyzed the global burden of childhood and adolescent asthma from 1990 to 2023 using GBD 2023 data. Compared with 1990, mortality and DALY rates significantly declined in 2023, while prevalence and incidence rates continued to rise. Future projections indicate an overall declining trend in the disease burden of childhood and adolescent asthma. These findings provide crucial evidence for developing differentiated and precision-based global asthma prevention and control strategies.

Geographic disparities represent a key finding of this study. The age-standardized prevalence in Australasia (5,994.4 per 100,000) was 8.2 times higher than in South Asia (731.9 per 100,000), demonstrating marked regional variations. Notably, HDI showed a positive correlation with asthma prevalence (Spearman R=0.244, P=0.001), indicating that socioeconomic development level correlates positively with asthma prevalence. This positive association contrasts with typical epidemiological patterns of most diseases, suggesting that asthma etiology may be closely linked to specific factors in socioeconomic development. Reduced early microbial exposure leads to abnormal immune system development, increasing the risk of allergic diseases [9]. Immunologically, decreased early childhood infections in developed countries result in insufficient Th1 immune responses and relative Th2 dominance, promoting IgE production and eosinophil activation. Concurrently, regulatory T cell (Treg) dysfunction impairs the suppression of excessive immune responses [10]. Microbiome research further supports this mechanism, with cesarean delivery and antibiotic use altering gut microbiota composition, affecting short-chain fatty acid production, and subsequently disrupting immune tolerance establishment [11, 12]. Additionally, environmental factors in developed countries, including urbanization, indoor lifestyles, air pollution, and increased allergen exposure, may synergistically promote asthma development [13]. Epigenetic studies reveal that environmental factors can influence asthma susceptibility gene expression through DNA methylation and histone modifications; these epigenetic modifications may be transgenerationally inherited, partially explaining the sustained upward trend in asthma incidence[14]. Despite higher prevalence in developed regions, mortality rates are significantly lower, with HDI showing a strong negative correlation with mortality (Spearman R=-0.837, *P*<0.001), reflecting the crucial role of healthcare accessibility and quality in improving asthma outcomes. This inverse relationship between prevalence and mortality has important implications for public health strategy development: developing countries should prioritize improving healthcare service quality to reduce mortality, while developed countries need to strengthen primary prevention through environmental and lifestyle interventions to control incidence. This geographic heterogeneity indicates that global asthma prevention and control require differentiated strategies based on regional epidemiological characteristics.

This study found that all epidemiological indicators were higher in males during childhood but showed significant reversal during adolescence (15-19 years), with females surpassing males. This phenomenon involves complex bio-psycho-social mechanism networks. At the molecular level, estrogen influences asthma pathogenesis through immune pathway regulation (such as promoting Th2 differentiation and enhancing mast cell degranulation) [15, 16]. In contrast, testosterone exhibits immunosuppressive effects, inhibiting IgE production and eosinophil infiltration [17]. Furthermore, factors including smaller airway caliber in females, menstrual cycle-related airway inflammation fluctuations, and increased sensitivity to environmental stimuli may promote asthma development [18]. Psychosocial factors, such as higher rates of anxiety and depression in adolescent females, may also affect asthma control [19]. Notably, prediction models indicate that female mortality and DALY rates will show an increasing trend by 2038, a trend potentially associated with modern lifestyle changes including rising smoking rates among adolescent females, pubertal stress, and hormonal contraceptive use [20]. These findings emphasize the necessity of developing sex-specific interventions, particularly asthma management strategies for adolescent females.

In addition to these long-term epidemiological trends, recent global public health events have significantly impacted childhood asthma epidemiological characteristics. The COVID-19 pandemic, as a major event at the study period’s end (2020-2023), provides a unique perspective for understanding the acute impact of environmental factors on asthma burden. Short-term effects manifested as protective factors: preventive measures including mask-wearing, social distancing, and home isolation significantly reduced respiratory pathogen transmission, thereby decreasing infection-triggered acute asthma exacerbations, with studies reporting a 30-50% reduction in emergency department visits [21]. However, long-term pandemic effects warrant attention. First, emerging long COVID syndrome is associated with persistent respiratory symptoms, with 12.7% of COVID-19-affected children showing sustained airway inflammation, potentially exacerbating pre-existing asthma[22]. Second, structural impacts on healthcare systems cannot be ignored: disruption of routine asthma clinic services compromised disease surveillance and management continuity, with medication adherence declining by 23%[23]. Third, altered environmental exposure patterns, with extended home confinement increasing cumulative exposure to indoor allergens (dust mites, mold, pet dander). Fourth, pandemic-related psychological stress, including social isolation, academic disruption, and family pressure, may influence asthma control through neuroimmune mechanisms[24]. While telemedicine partially compensated for reduced in-person consultations as an emergency measure, its effectiveness, accessibility, and equity in long-term childhood asthma management require systematic evaluation[25].

The BAPC prediction model shows that while the overall disease burden of childhood and adolescent asthma will exhibit a downward trend, female mortality and DALY rates are projected to show slight upward trends, indicating unique health challenges facing female children and adolescents. This prediction aligns with the observed pubertal sex-pattern reversal, reflecting the complex interaction of biological, environmental, and social factors differentially affecting populations by sex. Based on these epidemiological characteristics and future trend predictions, addressing current challenges in asthma prevention and control requires stratified clinical and public health interventions. First, high-risk population screening and early identification strategies should be implemented. Given the high incidence in the 5-9 age group, systematic screening using validated tools such as the Childhood Asthma Control Test (C-ACT) is recommended for high-risk children, with assessments every 6 months and timely specialist referral for those with C-ACT scores ≤19[26]. Concurrently, machine learning-based risk prediction models should be established, integrating genetic risk scores, environmental exposure data, and biomarkers (such as exhaled nitric oxide and blood eosinophils) for individualized risk assessment [27]. Second, addressing the predicted upward trend in female disease burden requires strengthened individualized management for adolescent females, including establishing menstrual cycle asthma diaries to identify hormone-related deterioration patterns, preventively adjusting controller medications premenstrually, providing psychological support and cognitive behavioral therapy, evaluating hormonal contraceptive effects on asthma, and enhancing education about e-cigarette hazards [28]. Third, considering the important role of environmental factors in asthma pathogenesis, implementing indoor air quality monitoring and improvement programs in schools, promoting low-allergen bedding and air purifier use, establishing extreme weather asthma warning systems, and strengthening regulation of emerging pollutants (microplastics, e-cigarettes) are recommended [29] [30].

This study has several limitations. First, GBD data quality varies across regions, with developing countries potentially experiencing significant underdiagnosis and misdiagnosis, possibly underestimating actual disease burden. Second, regional differences in diagnostic criteria may affect the accuracy of international comparisons. Third, GBD data cannot analyze asthma phenotypes (allergic, non-allergic, exercise-induced) and control levels, which significantly impact disease burden. Fourth, this ecological study design cannot establish individual-level causal relationships; observed associations may be subject to ecological fallacy. Fifth, the BAPC prediction model assumes historical trends will continue without considering potential breakthrough therapies or major policy changes that could affect future disease burden. Sixth, this study could not incorporate certain important environmental and social determinants, such as climate change and the implementation effects of air quality improvement measures, which may significantly influence future asthma trends. Finally, due to data limitations, this study could not analyze the combined impact of asthma comorbidities (allergic rhinitis, atopic dermatitis) on disease burden.

## 5 Conclusion

By analyzing GBD data from 1990-2023, this study comprehensively revealed the spatiotemporal distribution characteristics and trends of global childhood and adolescent asthma burden. Despite medical advances leading to significant declines in mortality and DALY rates, the continued rise in prevalence and incidence indicates that asthma remains a major public health challenge. Future research should focus on disease burden across different SDI regions and between sexes to provide more targeted evidence for global childhood and adolescent asthma prevention and control.

## Data Availability

The data analyzed in this study are publicly available from the Global Burden of Disease Study 2023 (GBD 2023) via the GBD Results Tool, which can be accessed at: http://ghdx.healthdata.org/gbd-results-tool. No additional data were collected by the authors.

## Author Contributions

EHY conceived, designed and supervised the study. HYY and WT collected and processed the data, and SRH and CXY contributed to drafting the manuscript. FD validated the findings. YY and ZXW provided critical feedback and helped shape the research. EHY and SRH revised the manuscript. All authors discussed the results and approved the final manuscript.

## Acknowledgments

We are grateful for the work of the Global Burden of Disease study 2023 collaborators.

## Availability of data

The data used in this study came from a public database that everyone can access through the link provided in this article (https://vizhub.healthdata.org/gbd-results/).

## Conflicts of Interest

The authors declare no conflicts of interest related to this study.

## Funding

The author(s) received no financial support for the research, authorship, and/or publication of this article.

## Notes

### Competing Interest Statement

The authors have declared no competing interest.

### Funding Statement

The author(s) received no specific funding for this work.

### Author Declarations

Ethical approval was not required for this study. This research is a secondary analysis of publicly available, aggregate-level, de-identified data from the Global Burden of Disease 2023 (GBD 2023) study, which does not involve human subjects, primary data collection, or identifiable personal information. Patient/participant consent is not applicable.

